# HIV-DRIVES: HIV Drug Resistance Identification, Variant Evaluation, & Surveillance Pipeline

**DOI:** 10.1101/2023.09.30.23296350

**Authors:** Stephen Kanyerezi, Ivan Sserwadda, Aloysius Ssemaganda, Julius Seruyange, Alisen Ayitewala, Hellen Oundo, Wilson Tenywa, Brian Kagurusi, Godwin Tusabe, Stacy Were, Isaac Ssewanyana, Susan Nabadda, Maria Magdalene Namaganda, Gerald Mboowa

**Affiliations:** Department of Immunology and Molecular Biology, School of Biomedical Sciences, College of Health Sciences, Makerere University, P.O Box 7072, Kampala, Uganda; The African Center of Excellence in Bioinformatics and Data-Intensive Sciences, the Infectious Diseases Institute, College of Health Sciences, Makerere University, P.O Box 22418, Kampala, Uganda; National Health Laboratories and Diagnostics Services, Central Public Health Laboratories, Ministry of Health, P.O Box 7272, Kampala, Uganda; Department of Medical Microbiology and Infectious Diseases, University of Manitoba, Winnipeg, MB, Canada; Africa Centres for Disease Control and Prevention, African Union Commission, Roosevelt Street, P.O. Box 3243, W21 K19, Addis Ababa, Ethiopia; Amsterdam Institute for Global Health and Development (AIGHD), Department of Global Health, Academic Medical Center, Amsterdam, The Netherlands

**Author notes:** To whom correspondence should be addressed: Gerald Mboowa. These authors contributed equally and share first authorship (Stephen Kanyerezi, Ivan Sserwadda).

**Keywords:** Bioinformatics, HIV drug resistance, Mutations, Next-generation sequencing

## Abstract

The global prevalence of resistance to the Human Immunodeficiency Virus (HIV) combined antiretroviral therapy (cART) emphasizes the need to continuous monitoring to better understand the dynamics of drug-resistant mutations to guide treatment optimization and patient management as well as check the spread of resistant viral strains. We have recently, integrated next-generation sequencing (NGS) into routine HIV drug resistance (HIVDR) monitoring, with key challenges in the bioinformatic analysis and interpretation of the complex data generated while ensuring data security and privacy of patient information. To address these challenges, here, we present HIV-DRIVES (HIV Drug Resistance Identification, Variant Evaluation, and Surveillance), an NGS-HIVDR bioinformatics pipeline that has been developed and validated using Illumina short-reads, FASTA, and sanger ab1.seq files.

**Availability and implementation:** HIV-DRIVES source codes and operation manual freely available at https://github.com/MicroBioGenoHub/HIV-DRIVES.

## Introduction

The rise of drug-resistant strains of the Human Immunodeficiency Virus (HIV) presents a formidable challenge in effectively managing HIV infections [1]. Precise identification and evaluation of HIV drug resistance (HIVDR) mutations are essential for guiding appropriate treatment decisions and devising effective therapeutic strategies [2]. Consequently, the demand for advanced bioinformatics pipelines that seamlessly integrate drug resistance identification, variant evaluation, and surveillance has become increasingly evident.

Traditionally, HIVDR profiling relied on tools such as the HIVdb program, a web-based program hosted by the Stanford University in California, along with RECall, and HyDRA, following the sequencing process [3]–[6]. While these tools have been invaluable in shedding light on drug resistance mutations, they exhibit limitations concerning data protection, transmission and automate the analysis. With web-based tools, concerns arise over the potential compromise of patient privacy due to the transmission of patient data over networks beyond countries borders. In contrast, certain command line tools conduct analyses in fragments, lacking the ability to offer a comprehensive end-to-end analysis and provide easily interpretable portable clinically actionable results. Additionally, these tools may not support analysis of genomic data from various sequencing platforms.

To address these challenges head-on, we introduce the HIV-DRIVES (HIV Drug Resistance Identification, Variant Evaluation, and Surveillance) bioinformatics pipeline. This high-level analytical pipeline has been purposefully designed to overcome the limitations inherent in traditional HIV drug resistance profiling methods. HIV-DRIVES offers a seamless and efficient approach for the detection, evaluation, and monitoring of HIV drug resistance mutations, harnessing the capabilities of advanced sequencing data and computational techniques. The development of HIV-DRIVES has been motivated by the goal of enhancing both patient care and public health, capitalizing on the potential of Next-Generation Sequencing (NGS) technologies.

## Implementation

### Data generation

The pipeline was tested on both data archived and generated by the National Genomics Reference Laboratory housed at the Central Public Health Laboratories. For newly generated data from the National Genomics Reference Laboratory, RNA was extracted using the QIAamp viral RNA extraction kit following the in-house customized protocol. The extracted RNA was reverse transcribed to cDNA which was later amplified using respective primer sets to generate amplicons. Quality assessment of the amplicons was performed using gel electrophoresis. Library preparation for the good quality amplicons was performed using the illumina DNA prep kit. To assess the quality of prepared libraries, DNA quantification and normalization using the qubit4 Fluorometer and library size estimation using agilent bioanalyzer were done. The libraries were loaded onto both the Illumina MiSeq and iSeq platforms for sequencing.

### Pipeline architecture

HIV-DRIVES is a tool designed with 3 programming languages that include, shell script, R, and Perl. It was compiled and tested on Ubuntu 18.0.4 LTS (Bionic Beaver). HIV-DRIVES was put together using different packages that include; trim_galore, bowtie2, samtools, quasitools, and sierra-local [6]–[10]. All these packages and their dependencies are housed in the HIV-DRIVES conda environment so that they do not interfere with already existing programs. The help message of the HIV-DRIVES was adapted from the rMAP help message and edited to suite the context of the HIV-DRIVES [11]. The full list of the dependent packages is provided in the table below.

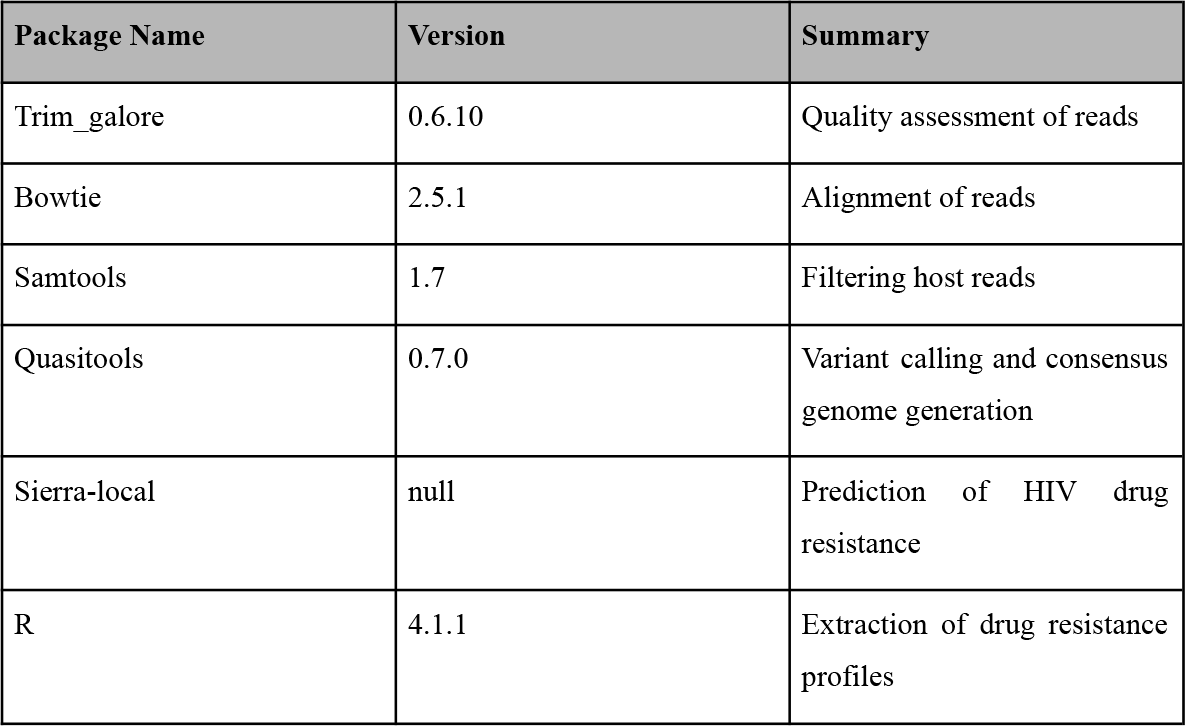

### Overview of the HIV-DRIVES pipeline workflow

HIV-DRIVES is designed to perform HIV drug resistance profiling and amino acid mutation detection of NGS data from illumina platforms, Sanger sequence data in *ab1*.seq format, and FASTA files. Given input data as FASTQ files, the tool uses trim_galore to filter out reads at a threshold phred score of *q*28. The remaining reads are aligned to the host genome (GRCh38) using bowtie2 to separate host and viral reads using samtools. The resultant viral reads are subjected to HyDRA from quasitools to generate a consensus genome and detect amino acid variants. The resultant consensus genome is subjected to sierra-local to predict the drug resistance within the genome. The output *json* file from sierra-local and amino acid mutations from HyDRA are interrogated using customized R, Perl, and Bash code to match the drug resistance profiles and their corresponding mutations plus comments which are finally output in a *pdf* file. In the *pdf* file, the resistance profiles are color coded. For Sanger and FASTA files, the tool uses sierra-local to determine the drug resistance with in the genome.

The output *json* file from Sierra-local interrogated using customized R, Perl, and Bash code to get the resistance profiles and their corresponding drug resistance mutations and comments which are finally output in a *pdf* file. The systematic flow of the pipeline is shown in the image below.

**Figure 1.**
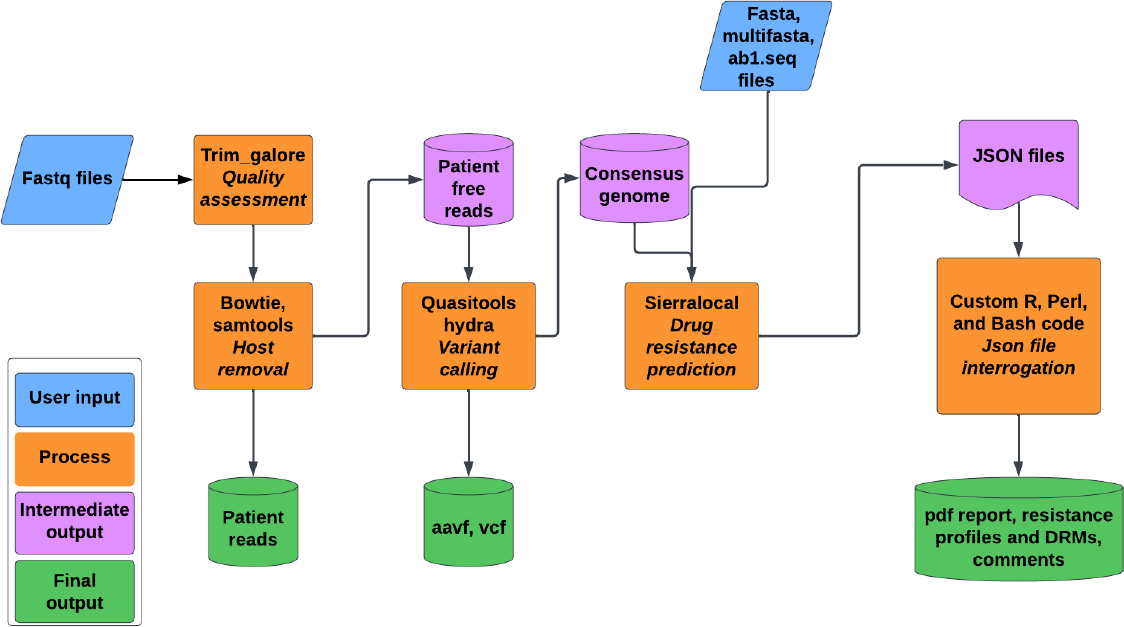
The systematic flow of the HIV-DRIVES pipeline

HIV-DRIVES can be run in 3 different modes, namely, all, varcall, and resistance. If one has got FASTQ files and would like to get resistance profiles at the end of the run, then they will need to turn on all mode and the corresponding options. If one has got FASTQ files and they only want to get the amino acid mutations and a consensus genome FASTA file, they will need to turn on the varcall mode and the corresponding options. If one has got a consensus genome FASTA file and would like to get resistance profiles at the end of the run, then they will need to turn on the resistance mode and the corresponding options. The resistance mode works for those with a consensus file in both FASTA and multifasta format, and those with a sanger output file in the format of *ab1*.seq. For all scenarios that require resistance profiling, the pipeline starts by updating the HIVDB resistance algorithm. The procedure of running all these modes is described in the core pipeline features section below.

### Core pipeline features

The core parameters of HIV-DRIVES are dependent on which mode the user is interested in running which is dependent on one’s needs, but the output directory is a mandatory parameter for all the modes. Here, we describe how to run the pipeline with different modes and the corresponding needs.

### All

The “all” mode is to be run when one has FASTQ files that are either paired or single ended and they want to get resistance profiles at the end.

For paired reads, below is how to run the tool;

HIV-DRIVES -o < output directory to be created > -f < path to the forward read > -r < path to the reverse read > --all true

For single ended reads, here is how to run the tool;

HIV-DRIVES -o < output directory to be created > --single-end true --se < path to the single ended read > --all true

### Varcall

The varcall mode is to be run when one has FASTQ files that are either paired or single ended and they only want amino acid mutations and consensus genome files generated at the end.

For paired reads, below is how to run the tool

HIV-DRIVES -o < output directory to be created > -f < path to the forward read > -r < path to the reverse read > --varcall true

For single ended reads, below is how to run the tool;

HIV-DRIVES -o < output directory to be created > --single-end true --se < path to the single ended read > --varcall true

### Resistance

Resistance mode is run when one has either the sanger *ab1*.seq file format or a FASTA file. It also supports a multifasta file. If one has a sanger file, below is how to run the tool

HIV-DRIVES -o < output directory to be created > --resistance true --sanger < path to the *ab1*.seq file >

If one has a FASTA file, below is how to run the tool

HIV-DRIVES -o < output directory to be created > --resistance true --consensus < path to the fasta file >

For all the modes, the resultant *pdf* files are output in the results directory under the output directory. The results directory also contains *aavf, vcf, resistance profile in csv*, drug resistance mutations corresponding to the drug classes, and the comments of mutations in *csv* files. For all and varcall modes, the output directory has patient_free_reads and patient_reads directories in which the viral reads and patient reads are found respectively. In cases of data sharing, the owner of the data can share the viral reads with confidence that patient genomic data is not shared.

### Testing and Validation

The pipeline was tested on samples sequenced on MiSeq and iSeq sequencing platforms at the National Genomics Reference Laboratory housed at the Central Public Health Laboratories, illumina and Sanger samples from https://f1000research.com/articles/11-901, as well as protease and integrase sequences from the Stanford University HIV drug resistance database (https://hivdb.stanford.edu/hivdb/by-sequences/) [3], [4], [12]. HIV-DRIVES was benchmarked with the classical Stanford University HIVdb program (https://hivdb.stanford.edu/hivdb/by-sequences/) with the resistance profiles and drug resistant mutations that contribute to the resistance as the metrics of comparison [3], [4]. For each sample, we obtained drug resistance profiles for antiretroviral drugs of the following drug classes; protease inhibitors (PIs), non-nucleoside reverse transcriptase inhibitors (NNRTIs), nucleoside reverse transcriptase inhibitors (NRTIs) and integrase strand transfer inhibitors (INIs) with the drug resistance mutations contributing to the resistances. Additionally, the corresponding comments to the mutations were extracted. For all the metrics, there was 100% concordance with the Stanford University HIVdb program. The validated reports for the HIV-DRIVES compared with stanford can be found at https://github.com/MicroBioGenoHub/HIV-DRIVES-validation-reports

## Conclusion

In summary, the HIV-DRIVES (HIV Drug Resistance Identification, Variant Evaluation, and Surveillance) bioinformatics pipeline stands as a potent and pioneering instrument within the realm of HIV drug resistance surveillance and treatment. This pipeline effectively harnesses state-of-the-art sequencing data and computational methodologies to identify, assess, and track mutations associated with HIV drug resistance. Consequently, it bolsters our capacity to comprehend and counteract HIV drug resistance, ultimately contributing to the development of more efficient treatment strategies tailored to public health needs and the enhancement of patient care. The incorporation of this pipeline into the infrastructure of the Central Public Health Laboratory (CPHL) in Uganda for routine HIVDR care not only solidifies its relevance in public health but also underscores its potential for adoption in similar institutions worldwide.

## Availability and future directions

The source code is available on GitHub under GPL3 license at https://github.com/MicroBioGenoHub/HIV-DRIVES. Questions and issues can be sent to ““.

Future directions of HIV-DRIVES consist of implementation within Singularity, Docker and Nextflow platform containers as well as the integration of further enhancements in terms of scalability and usability.

## Ethics declarations

This study data was obtained following permission to develop the HIV Drug Resistance Identification, Variant Evaluation, & Surveillance Pipeline (HIV-DRIVES) to be used by institutions within Uganda. The need for approval was waived by the research ethics committee of Uganda National Health Laboratory Services, Kampala Uganda.

## Data availability statement

The data generated from the National Genomics Reference Laboratory housed at the Central Public Health Laboratories is available upon request.

## Acknowledgments

We would like to express our heartfelt gratitude to the developers of Sierra-local and Quasitools for their invaluable contributions to the development of HIV-DRIVES. Their exceptional tools have played a pivotal role in enhancing the functionality and efficiency of our work, and we are sincerely thankful for their dedication and expertise.

## Funding

S.K is grateful to the Eastern Africa Network of Bioinformatics Training (EANBiT) project team for their support. The views and opinions of the authors expressed herein do not necessarily state or reflect those of EANBiT

S.K is also a Research Fellow under the African Doctoral Dissertation Research Fellowship (ADDRF) award offered by the African Population and Health Research Center (APHRC) and funded by the Bill and Melinda Gates Foundation through a project grant to APHRC

S.K, G.M. and I.S acknowledge the EDCTP2 career development grant which supports the Pathogen detection in HIV-infected children and adolescents with non-malarial febrile illnesses using the metagenomic next-generation sequencing approach in Uganda (PHICAMS) project which is part of the EDCTP2 programme from the European Union (Grant number TMA2020CDF-3159). The views and opinions of the authors expressed herein do not necessarily state or reflect those of EDCTP.

G.M is equally grateful for the support of the NIH Common Fund, through the OD/Office of Strategic Coordination (OSC) and the Fogarty International Center (FIC), NIH award number U2RTW010672. Its contents are solely the responsibility of the authors and do not necessarily represent the official views of the supporting offices”.

I.S is a PhD fellow at the Amsterdam Institute of Global health and Development (AIGHD) funded by the EDCTP Scholarship program through the CAGE-TB Project. Part of his work is also funded by the Public Health Alliance for Genomic Epidemiology (PHA4GE) grant number (INV-038071) supporting pathogen genomics data standards and bioinformatics interoperability.

## Competing interests

The authors have declared that no competing interests exist.

## Notes

### Competing Interest Statement

The authors have declared no competing interest.

### Funding Statement

This study did not receive any funding

### Author Declarations

Research Ethics Committee of the National Health Laboratory Services, Kampala Uganda waived ethical approval for this work

